# Forecasting admissions in psychiatric hospitals before and during Covid-19

**DOI:** 10.1101/2021.07.16.21260200

**Authors:** Jan Wolff, Ansgar Klimke, Michael Marschollek, Tim Kacprowski

## Abstract

**Introduction:** The COVID-19 pandemic has strong effects on most health care systems and individual services providers. Forecasting of admissions can help for the efficient organisation of hospital care. We aimed to forecast the number of admissions to psychiatric hospitals before and during the COVID-19 pandemic and we compared the performance of machine learning models and time series models. This would eventually allow to support timely resource allocation for optimal treatment of patients.

**Methods:** We used admission data from 9 psychiatric hospitals in Germany between 2017 and 2020. We compared machine learning models with time series models in weekly, monthly and yearly forecasting before and during the COVID-19 pandemic. Our models were trained and validated with data from the first two years and tested in prospectively sliding time-windows in the last two years.

**Results:** A total of 90,686 admissions were analysed. The models explained up to 90% of variance in hospital admissions in 2019 and 75% in 2020 with the effects of the COVID-19 pandemic. The best models substantially outperformed a one-step seasonal naïve forecast (seasonal mean absolute scaled error (sMASE) 2019: 0.59, 2020: 0.76). The best model in 2019 was a machine learning model (elastic net, mean absolute error (MAE): 7.25). The best model in 2020 was a time series model (exponential smoothing state space model with Box-Cox transformation, ARMA errors and trend and seasonal components, MAE: 10.44), which adjusted more quickly to the shock effects of the COVID-19 pandemic. Models forecasting admissions one week in advance did not perform better than monthly and yearly models in 2019 but they did in 2020. The most important features for the machine learning models were calendrical variables.

**Conclusion:** Model performance did not vary much between different modelling approaches before the COVID-19 pandemic and established forecasts were substantially better than one-step seasonal naïve forecasts. However, weekly time series models adjusted quicker to the COVID-19 related shock effects. In practice, different forecast horizons could be used simultaneously to allow both early planning and quick adjustments to external effects.

## I. Introduction

**H**ealth care systems need to balance potentially unlimited demand for services with scarce health care resources [1]. This balance might become even more difficult in the future due to an aging population that leads to increased demand for health care services and reduced medical work force [2]. Hospitals often consume a large part of total health care budgets [3]. A possible way to mitigate resource constraints in hospitals is the use of modern technologies to make services more cost-effective [4]. For instance, the increasing availability of data allows to support medical decision making in hospitals with information derived from machine learning algorithms [5, 6].

Efficient resource allocation in hospitals requires the management of volatile demand and available resources. This management is more critical in hospitals than in other areas since lack of timely and sufficient services can lead to negative patient outcomes [7]. Insufficient staff can lead to increased morbidity and mortality [8, 9]. Due to the shortage of trained medical staff in many health care systems, securing sufficient medical staff to permanently meet patient needs becomes a critical objective [10]. Inpatient mental health care is more staff intensive than other medical disciplines due to the personal nature of many interventions [11, 12].

The Covid-19 pandemic has strong effects on most health care systems and individual service providers [13]. A sudden surge in patients requiring intensive respiratory care and the possible shortage of ICU capacities led to political supply side interventions in many health care systems [14]. In Hesse, Germany, somatic and psychiatric hospitals were required to restrict new admissions to urgent care cases and avoid elective patients from 16th March 2020. Furthermore, new hospital hygiene regulations and the requirement to quarantine patients reduced hospital capacities.

Forecasting of admissions can help for the efficient organisation of hospital care and for the adjustment of resources to sudden changes in patient volumes. We aimed to forecast the number of admissions to psychiatric hospitals before and during the COVID-19 pandemic and we compared the performance of machine learning models and time series models.

## II. Methods

### A. Data

We included all inpatient admissions from 01. January 2017 to 31. December 2020 to nine hospitals in Hesse, Germany. These hospitals are part of a common service provider and account for about half of all inpatient mental health care in the state of Hesse. Aggregated admission numbers per day were obtained from the hospital administrations and did not contain individual patient data. Returns after planned interruptions, such as home leave, were excluded. Multiple separate admissions of the same patient were counted individually. Admissions to the departments of child and adolescent psychiatry and admissions to the departments for psychosomatic medicine were excluded.

We obtained weather and climate data from the Climate Data Centre of Germany’s National Meteorological Service [15]. We used the gtrendsR package to query Google trend data for Hesse, Germany [16]. School holidays and public holidays were obtained from publicly available calendars.

### B. Analyses

We used machine learning and time series models to predict the number of hospital admissions for each day of year 2019 and 2020. The machine learning models were a) gradient boosting with trees (XGB) [17], b) support vector machines (SVM) [18] and c) elastic nets [19]. The time series models were a) exponential smoothing state space models (ETS) [20], b) exponential smoothing state space models with screening for Box-Cox transformation, ARMA errors and trend and seasonal components (TBATS) [21] and c) additive models with non-linear trends fitted by seasonal effects (PROPHET) [22]. We compared models forecasting a week in advance, a month in advance and a whole year one week in advance.

### C. Features

Our machine learning model used calendrical variables, climate and weather data, google trend data, Fourier terms and lagged number of admissions as features. The calendrical features were day of week, weekend, public holiday, school holiday, quarter of the year, month of the year, bridge days, i.e. days between a public holiday and the weekend and the end of the year, i.e. the days between Christmas and new year’s eve. The climate and weather data were wind speed, cloudiness, air pressure, precipitation depth and type, duration of sunshine, snow height, air temperature and humidity. Since the weather of future days was unknown at the point of prediction we used lagged values, i.e. the weekly model used the weather 7 days before the predicted day and the monthly models used the weather data 28 days before the predicted day. We did not use weather data for the yearly model.

We used the following keywords in google trend data: depression, sadness, suicide, mania, fear, panic, addiction, dependence, alcohol, drugs, schizophrenia, psychosis and hallucinations. As for the weather data, we used lagged values of google trend data. The weekly models used the number of admissions 14 days before the predicted day, because the number of admissions was not known yet on day 7 before prediction, as additional feature and the monthly model used these values with a lag of 35. Our time series models did not use feature variables.

### D. Training and testing

We used prospectively sliding time windows to validate (2018) and test (2019 and 2020) model performance. The final weekly models predicted each day of one full week of hospital admissions seven days in advance. We tested one model for each week and study site in 2019 and 2020, thereby incrementally prolonging the training period and forwarding the 7-day testing period each by one week. The monthly models each predicted 28 days of hospital admissions in advance and the incremental slides were 28 days. In the yearly models, we predicted the whole year of 2019 and 2020 each one week before the years started.

We compared model performance with the Root-Mean-Squared-Error (RMSE), the R2, the Mean Absolute Error (MAE) and a seasonal Mean Absolute Scaled Error (sMASE). The sMASE [23] was calculated by dividing the MAE of our weekly, monthly and yearly forecasts by the MAE derived from a naïve forecast based on the number of admissions observed 14 days, 35 days and 364 days before the predicted day, respectively.

## III. Results

The number of admissions showed a relatively strong weekly seasonality and a yearly seasonality. Figure 1 provides the results of a multiple seasonal decomposition of the number of daily admissions by loess [24]. There was no strong trend in admission numbers during the first three years, until the commencement of the Corona hospital regulation on March 16th had a clear negative effect on the number of admissions.

**Fig. 1.**
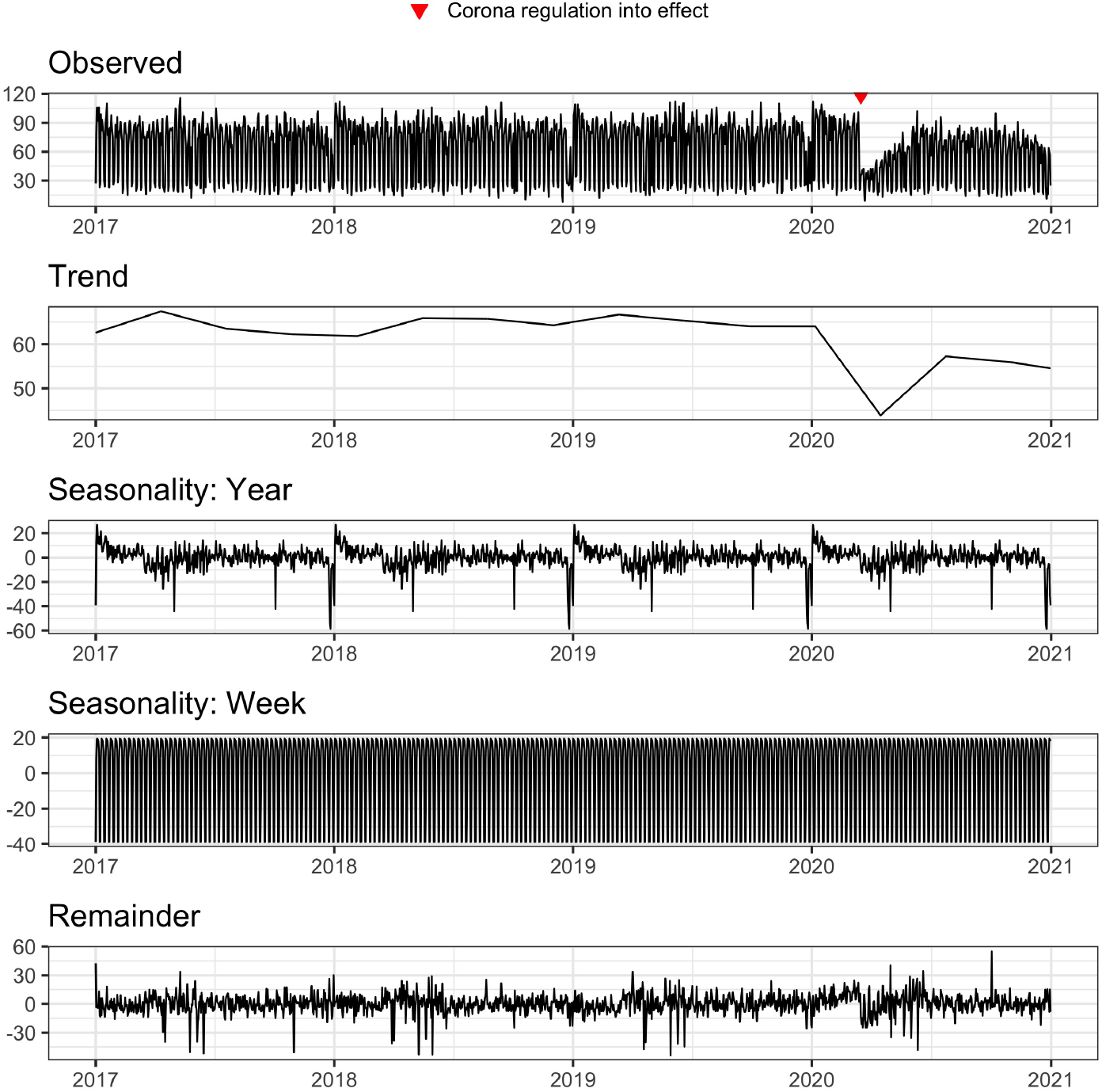
Multiple seasonal decomposition by loess. The y-axes are scaled differently.

Table I shows the forecasting performance in 2019 and in 2020 at all study sites combined. The naïve seasonal forecasts were based on the number of admissions 14, 35 and 364 days before the predicted day for the weekly, monthly and yearly models, respectively. In absolute terms, the best model in 2019 was the weekly elastic net, which achieved a MAE of 7.25 days and an explained variance of 90%. Compared to a naïve forecast based on the number of admissions two weeks in advance, this model achieved a forecast improvement of 38% (sMASE=0.62). In absolute terms, the best model in 2020 was the weekly TBATS model. However, compared to the monthly possible naïve forecast, i.e. the number of admission 35 days in advance, the highest improvement was achieved with the monthly SVM.

**TABLE I.**
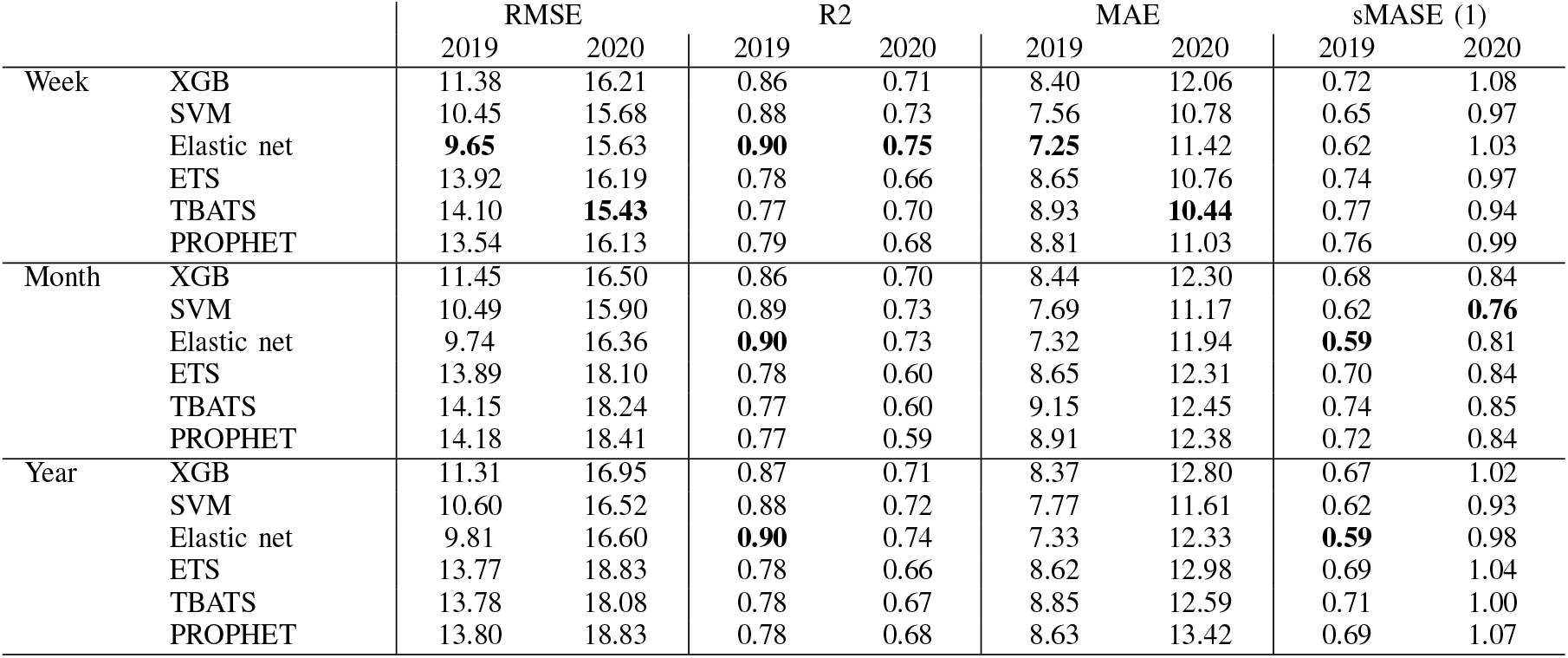
Forecasting performance in 2019 and in 2020. Best values per column are in boldface. RMSE= Root-Mean-Squared-Error, MAE= Mean Absolute Error, sMASE= Seasonal Mean Absolute Scaled Error. 1: The naïve seasonal forecasts were based on the number of admissions 14, 35 and 364 days before the predicted day for the weekly, monthly and yearly models, respectively.

The error accumulation in 2019 and 2020 at all study sites combined is shown in Figure 2. While model performance was relatively similar in 2019, errors diverged after commencement of the Corona hospital regulation on March 16th, 2020. Weekly time series models adjusted quicker to the new circumstances and accumulated less error until the end of year 2020.

**Fig. 2.**
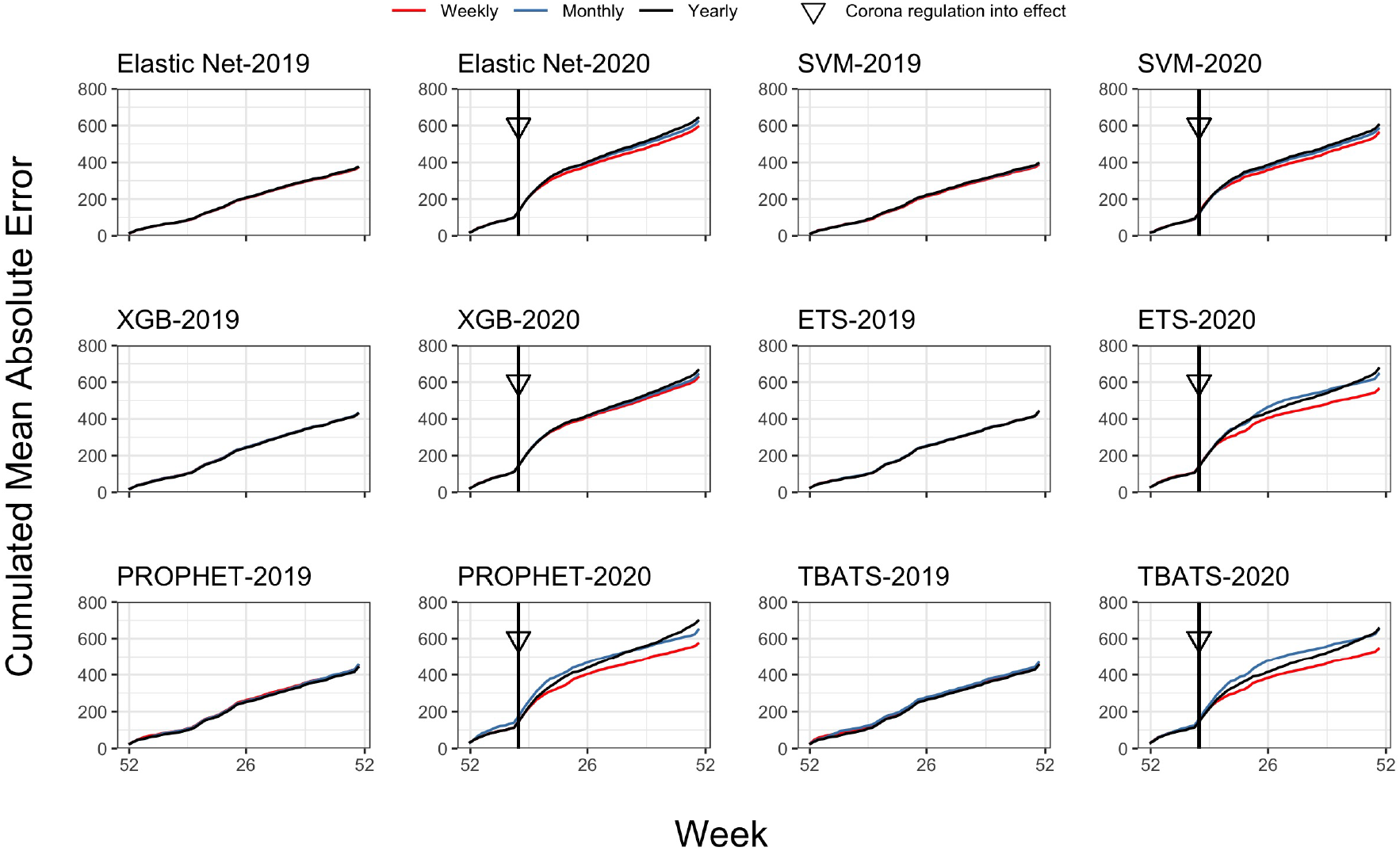
Cumulated mean absolute error in 2019 and 2020 by machine learning and time series models. XGB= Gradient boosting with trees, SVM= Support vector machines, ETS= Exponential smoothing state space models, TBATS= Exponential smoothing state space models with Box-Cox transformation, ARMA errors and trend and seasonal components, PROPHET= Additive models with non-linear trends fitted by seasonal effects.

The forecasting models showed variation in performance between study sites. Figure 3 shows differences in percentage errors between study sites per week derived from the overall best performing weekly machine learning and time-series models (see Table I), respectively. Both models performed similar in year 2019. However, the elastic net caused less error peaks, for instance at easter Monday and during Christmas time because it had these holidays as features. In contrast, the TBATS model adjusted quicker to the corona regulations and adjusted to the new level of admission numbers during the rest of 2020 better than the elastic net.

**Fig. 3.**
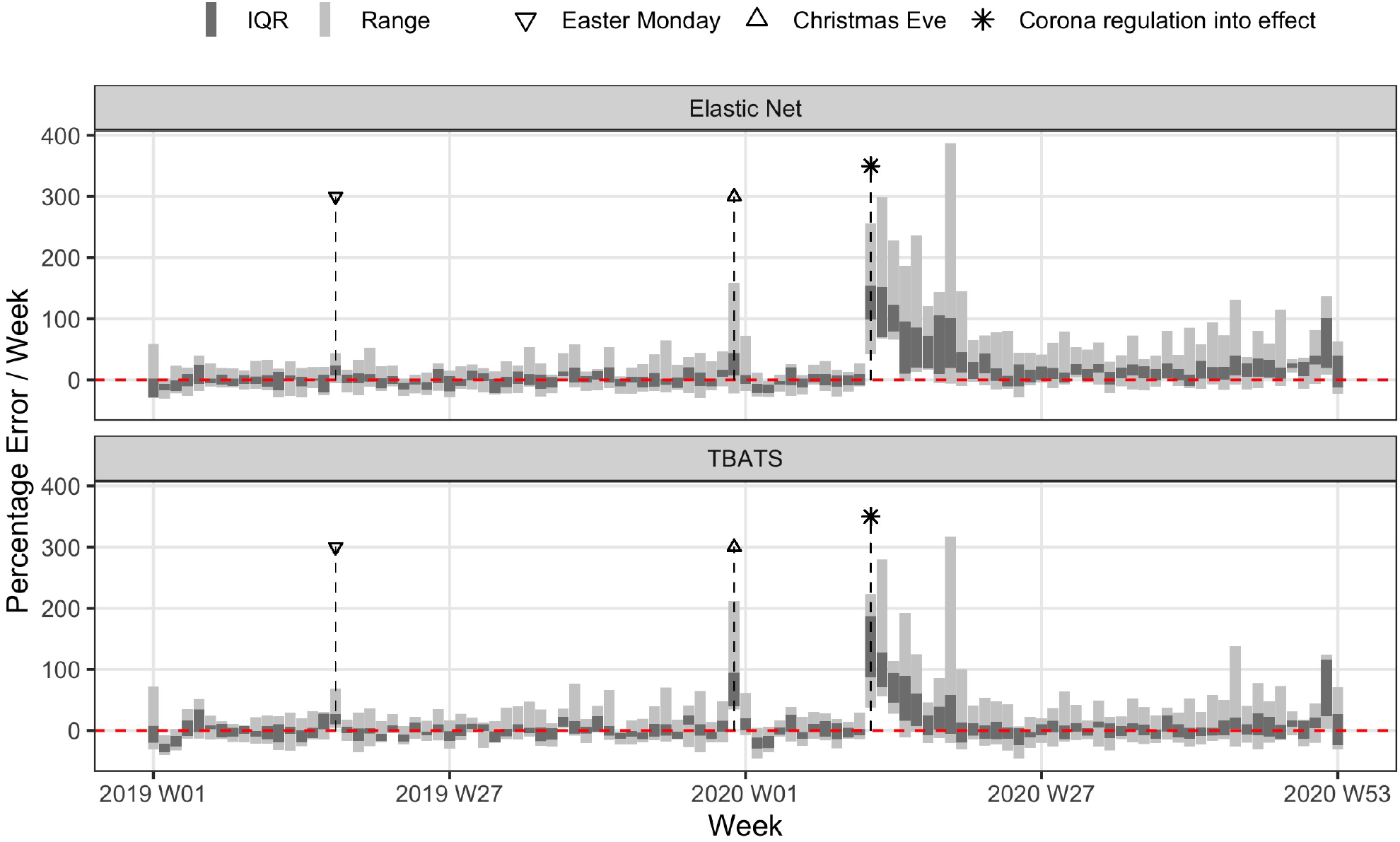
Variation of percentage error between study sites. TBATS= Exponential smoothing state space models with Box-Cox transformation, ARMA errors and trend and seasonal components, IQR= Interquartile range.

Figure 4 shows the top 25 feature variables ordered by their importance in forecasting the number of admissions with the elastic net, which was the best performing machine learning algorithm in our comparison. Variable importance represents the influence of each feature on the forecast performance relative to the other variables [25]. The strongest influence on forecast performance was found in calendrical variables.

**Fig. 4.**
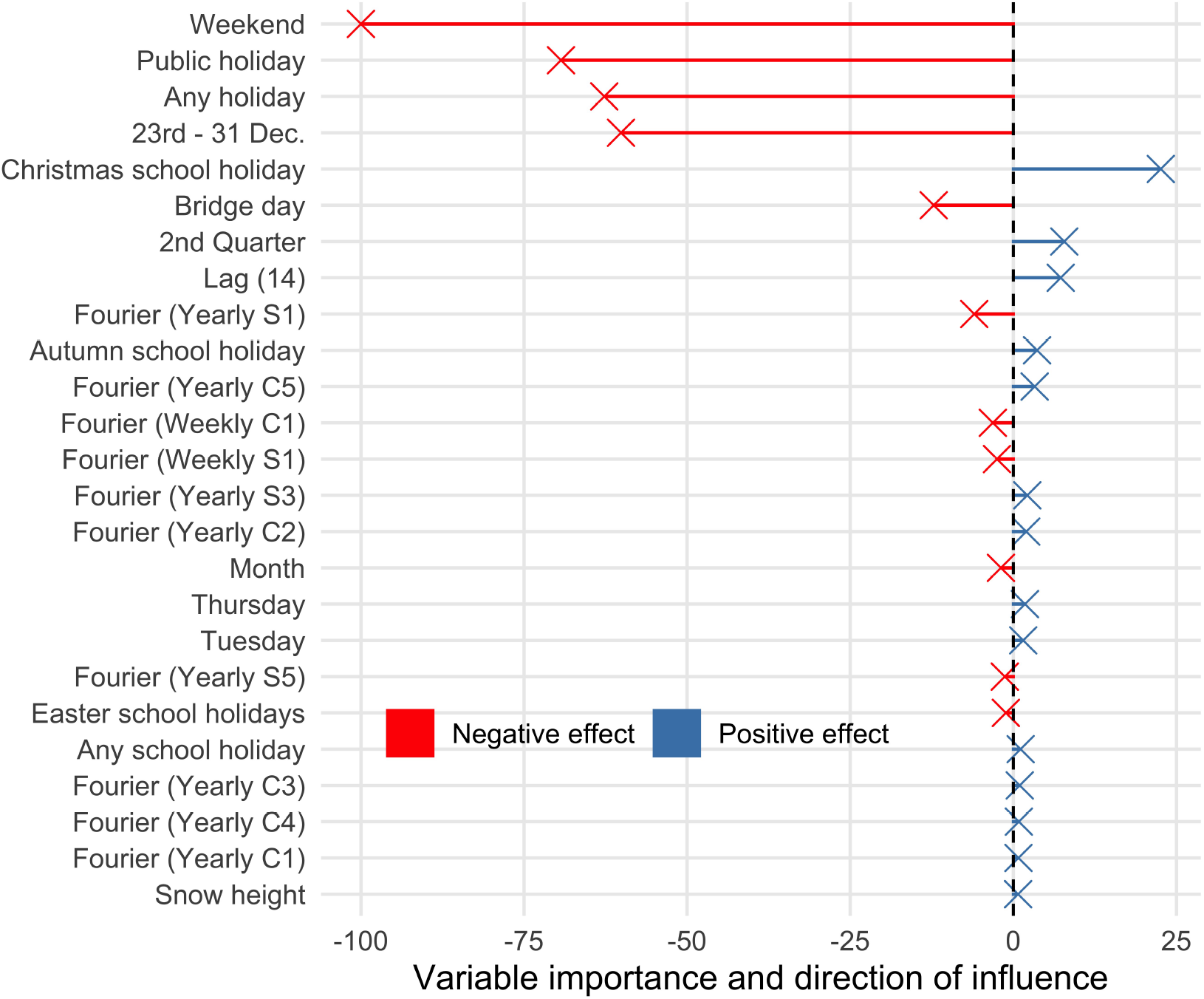
Variable importance of TOP 25 features in machine leaning models. Dec= December.

## IV. Discussion

### A. Key findings

We aimed to forecast the number of admissions to psychiatric hospitals before and during the COVID-19 pandemic and we compared the performance of machine learning models and time series models. This would eventually allow to support timely resource allocation for optimal treatment of patients. Model performance did not vary much between different modelling approaches before the COVID-19 pandemic. Established forecasts were substantially better than seasonal naïve forecasts. The most important features were calendrical variables that did not require short term adjustments in weekly and monthly models. However, weekly time series models adjusted quicker to the COVID-19 related shock effects than monthly and yearly models and the machine learning models.

### B. Strength and weaknesses

A strength of our study were the data of four years from nine hospitals representing about half of all inpatient psychiatric admissions in Hesse, Germany. This allowed both to give a representative picture of inpatient psychiatric care in Germany and to show how the forecasting approaches work at different study sites. Furthermore, it was possible to analyse the effect of sudden changes in hospital admissions to the performance of different modelling approaches due to the commencement of the Corona hospital regulation in March 2020.

A limitation of our study was the lack of data to differentiate between causes of reduced hospital admissions after the corona regulation came into effect in March 2020. The reduced admissions could have been a result of different supply side and demand side effects, such as avoidance of elective admissions, reduced capacities due to isolation and quarantine requirements and unwillingness of patients to enter hospitals during the Corona crisis. Another limitation of our study was its restriction to one large German provider of inpatient mental health care, which requires a lot of care when translating to different healthcare systems or different clinical settings.

### C. Comparison to previous research

Previous studies often focused on emergency departments [26] and there were no previous studies that analysed forecasting of psychiatric hospital admissions comparable to our study in scale and scope.

Vollmer *et al* 2021 predicted admission numbers in the emergency departments of two hospitals in London with data from 2011 to 2018 [27]. They compared machine learning models to more traditional time series models to make forecasts of admissions one, three and seven days in advance. The forecasts of different time horizons, i.e., one, three and seven days in advance, performed very similar. This is comparable to our findings of relatively similar results between weekly, monthly and yearly predictions, although at a different scale. In contrast to our study, lagged admissions from previous weeks were among the strongest predictors, probably related to the stronger increase and decrease of admission number levels during the study period at the different hospitals in comparison to our study. As in our study, Vollmer *et al* also found that calendrical variables were among the features with the strongest influence on forecasting performance. Weather and climate data and google search data had a relatively low influence on forecasting performance.

Similar results were found by Boutsioli *et al*, who used a simple OLS regression to forecast hospital admissions to the emergency departments of ten public hospitals in Greece [28]. They only used the calendrical variables weekend, summer holiday, public holiday and the participation in emergency care in their model and explained a relatively high variance in hospital admissions of up to 88%.

Jones *et al* forecasted the admission numbers at three emergency departments in the USA one, seven, fourteen, twenty-one and thirty days in advance [29]. They used autoregressive integrated moving average (ARIMA) models, time series regression, exponential smoothing, and artificial neural network models to predict admissions per day. They also found that admissions were characterised by yearly and weekly seasonality (see for comparison our Figure 1). As in our study, they found a relatively low improvement in forecasting performance in the shorter forecasting horizons in comparison to the longer horizons. Similar to our study and to the study of Vollmer *et al* [27], weather and climate had a relatively low influence on forecasting performance.

McCoy *et al* forecasted hospital discharge numbers at two academic medical centers in the USA [30]. They compared the performance of a PROPHET model with a seasonal ARIMA model and a one-step naïve seasonal forecast and compared monthly models to yearly models. The best performance was achieved by a PROPHET model. Comparable to our study, they also found relatively low to none improvement of forecasting accuracy in refitting their models monthly in comparison to yearly models.

## V. Conclusions

Accurate forecasting of hospital admissions can help for the efficient organisation of hospital care and for the adjustment of resources to sudden changes in patient volumes. We found a substantial improvement of forecasting accuracy in comparison to a seasonally adjusted naïve baseline forecast. Model performance did not vary much between different modelling approaches and different forecasting horizons before the COVID-19 pandemic. However, weekly time series models adjusted quicker to the COVID-19 related shock effects. In practice, different forecast horizons could be used simultaneously to allow both early planning and quick adjustments to external effects.

## Data Availability

Data can be provided upon reasonable request.

## Notes

### Competing Interest Statement

The authors have declared no competing interest.

### Funding Statement

We received no external funding.

### Author Declarations

The ethics committee of the Medical School Hannover confirmed that our study did not require ethical oversight.

